# EEG-guided early cessation of sedation and TTM in patients after cardiac arrest: a feasibility and safety study

**DOI:** 10.64898/2026.02.20.26345728

**Authors:** Marleen C. Tjepkema-Cloostermans, Albertus Beishuizen, Aart C. Strang, Hanneke M. Keijzer, Johan A. Telleman, Simone P. Smook, J. Wytze Vermeijden, Jeannette Hofmeijer, Michel J. A. M. van Putten

## Abstract

**Objective:** Despite substantial variability in the severity of post-anoxic encephalopathy, all comatose patients after cardiac arrest are usually treated according to the same standardized intensive care protocol, including sedation, mechanical ventilation, and targeted temperature management (TTM). We hypothesize that patients with a favourable EEG pattern (continuous EEG within 12 hours after cardiac arrest) may not benefit from prolonged sedation and TTM. We studied the feasibility and safety of early cessation of sedation and TTM in this subgroup.

**Methods:** We conducted a non-randomized, controlled intervention study including 40 adult patients admitted to the ICU with postanoxic encephalopathy after cardiac arrest and an early (*<* 12 hours) favourable EEG pattern. The control group received standard care with sedation and TTM for at least 24–48 hours, whereas the intervention group underwent early cessation of sedation and TTM as soon as possible after establishing a favourable EEG, followed by weaning from mechanical ventilation. The primary outcome was duration of mechanical ventilation. Secondary outcomes included ICU length of stay, total sedation time, number of ICU complications, and neurological outcomes at 3 and 6 months.

**Results:** Duration of mechanical ventilation was significantly shorter in the intervention than in the control group (median 12 vs 28 h, *p <* 0.001). Median ICU length of stay and median total sedation time were also reduced by more than 50% in the intervention group, from respectively 2.5 to 1.2 days (*p* = 0.001) and 27 to 12 h (*p <* 0.001). There was no increase in ICU complications in the intervention group. No statistically significant differences in neurological outcomes at 3 or 6 months were observed.

**Conclusion:** Early withdrawal of sedation is feasible and safe in patients with an early favourable EEG following cardiac arrest. The study was underpowered to detect possible differences in long-term neurological recovery.

**Significance:** Shortening sedation and mechanical ventilation is likely to result in direct reductions in healthcare costs and contribute to more appropriate care. Larger studies are needed to evaluate the impact on long-term neurological outcomes.

## 1. Introduction

Outcomes of comatose patients after cardiac arrest are highly heterogeneous. Approximately half of these patients never regain consciousness due to diffuse postanoxic encephalopathy [1, 2]. Among survivors, neurological recovery spans a broad spectrum, reflecting substantial interindividual variability in the severity of brain injury [3, 2]. Despite this heterogeneity, post–cardiac arrest care for patients who remain comatose after successful resuscitation is largely protocolized. Patients are routinely admitted to the intensive care unit (ICU), treated with sedation, and mechanically ventilated for at least 24–48 hours. In addition, body temperature is typically controlled using targeted temperature management (TTM) [1, 4]. However, the treatment remains uncertain, and potential benefits of sedation and hypothermia may be restricted to specific patient subgroups, depending on the initial electrocardiographic rhythm [5, 6] and the severity of encephalopathy [7, 8, 9].

A reliable bedside biomarker to assess encephalopathy severity is the electroencephalogram (EEG). EEG directly measures brain activity, and background patterns are robust predictors of long-term neurological outcome after cardiac arrest [3, 2, 10, 11]. Accordingly, continuous EEG monitoring has increasingly become routine and is incorporated in international prognostication guidelines [9]. A continuous background EEG pattern within 12 hours after cardiac arrest (“favourable EEG”) indicates transient or mild postanoxic encephalopathy and reliably predicts good neurological outcome [2, 10, 11]. We hypothesise that patients with a favourable early EEG do not benefit from prolonged deep sedation, TTM and mechanical ventilation.

In the current study, we investigated the feasibility and safety of EEG-guided early discontinuation of sedation and TTM in patients with a favourable EEG within 12 hours after cardiac arrest.

## 2. Methods

### 2.1. Study design and patient population

We conducted a prospective controlled interventional study with a sequential (before–after) design at two tertiary care hospitals in the Netherlands (Medisch Spectrum Twente and Rijnstate Hospital). Continuous EEG (cEEG) monitoring was part of routine care and was initiated as soon as possible after ICU admission.

Adult patients (≥ 18 years) who remained comatose after cardiac arrest (GCS ≤ 8) were screened for eligibility. Patients were included if the early EEG showed a favourable background pattern within 12 hours after cardiac arrest, defined as continuous normal-voltage background activity [3, 2]. Additional inclusion criteria were (i) clinical feasibility to discontinue sedative treatment within three hours after identification of the favourable EEG pattern and (ii) availability of written informed consent obtained through a deferred-consent procedure. Exclusion criteria were a known medical condition associated with a life expectancy of less than 6 months; a history of progressive neurological disease; a pre-admission GOS-E score of ≤ 4; non-neurological indications for continued sedation and/or mechanical ventilation (including cardiogenic shock or massive aspiration); and inability to complete follow-up due to logistical reasons.

During the first study phase, consecutive included patients received standard care (control cohort). After inclusion of 20 patients, both centres transitioned to the intervention phase. During this second phase, 20 consecutive included patients underwent early discontinuation of sedation and temperature management as soon as clinically feasible, followed by weaning from mechanical ventilation when appropriate.

Written informed consent was obtained using deferred-consent. This was requested from a legal representative as soon as possible after inclusion, but no later than 72 hours. In addition, all surviving patients were asked to provide written informed consent either during their hospital stay after regaining consciousness or during follow-up at 3 or 6 months. An independent Data and Safety Monitoring Board evaluated safety prior to the start of recruitment, after inclusion of 15 patients (75%) in the control arm, and after inclusion of 5 (25%) and 10 (50%) patients in the intervention arm. This was an investigator-initiated trial with no commercial involvement. The trial was registered in the ClinicalTrials.gov registry (NCT06048796), and approval was obtained from the Medical Research Ethics Committee (Medical research Ethics Committees United in Nieuwegein, study number NL84714.100.23).

### 2.2. ICU treatment

All patients received standard post–cardiac arrest ICU care, including sedation and mechanical ventilation. Temperature management followed the TTM-2 approach [1]: active cooling was initiated only in case of fever (≥ 37.8 °C) using a surface or intravascular device, targeting 37.5 °C. No active warming or cooling was provided when body temperature remained below 37.8 °C.

After inclusion, patients in the control cohort continued standard care with sedation, mechanical ventilation, and temperature management for at least 24 hours. In the intervention cohort, sedation and temperature management were discontinued as soon as feasible after detection of the favourable EEG, followed by weaning from mechanical ventilation when appropriate.

### 2.3. Baseline characteristics

Baseline characteristics collected included age, sex, location of cardiac arrest, presumed cause of cardiac arrest, first monitored electrocardiographic rhythm, time from cardiac arrest to initiation of basic life support, time from cardiac arrest to return of spontaneous circulation, and clinical characteristics at hospital admission. Admission characteristics included the Acute Physiology and Chronic Health Evaluation IV (APACHE IV) score, total Glasgow Coma Scale score, and the presence or absence of pupillary light reflexes.

### 2.4. Outcome parameters

The primary outcome was the duration of mechanical ventilation, expressed in hours. To further assess feasibility, secondary outcomes included length of ICU stay and total duration of sedation. Safety was evaluated by recording ICU-related complications, including autodetubation, death, pneumonia, sepsis, bleeding, delirium, cardiac arrhythmias, recurrent cardiac arrest, thrombocytopenia, and other serious adverse events (SAEs). In addition, the need for reintubation, resumption of sedation and administration of haloperidol were recorded as secondary outcome measures.

To assess long-term functional and cognitive recovery, follow-up evaluations were conducted by telephone at three and six months after admission by one of the researchers (MT-C) or a trained research nurse (EvV). During these interviews, neurological outcome was assessed using the Glasgow Outcome Scale–Extended (GOSE) and the Cerebral Performance Category (CPC) score, based on a standardised interview. Cognitive functioning was evaluated using the Telephone Montreal Cognitive Assessment (T-MoCA). In addition, mortality and the occurrence of serious adverse events (SAEs) after hospital discharge within six months were recorded. At the three-month follow-up, patients’ experiences with the deferred consent procedure were also assessed using a short questionnaire.

### 2.5. Statistical analyses

Baseline characteristics, the primary outcome, and all secondary outcomes were summarised descriptively using medians and interquartile ranges or proportions, as appropriate. Differences between the control and intervention groups were assessed using the Mann–Whitney U test for continuous variables and the Pearson chi-square test or Fisher’s exact test for categorical variables, as appropriate. All analyses were conducted according to the intention-to-treat principle.

## 3. Results

### 3.1. Baseline characteristics

Between March 2024 and May 2025, a total of 40 patients (26 at MST and 14 at Rijnstate) were successfully included, with 20 patients allocated to the control group and 20 to the intervention group. Baseline characteristics did not differ significantly between the control and intervention groups (Table 1). Figure 1 shows EEG fragments of two patients who were included in the study.

**Table 1:** Baseline characteristics of the study population.

| Characteristic | Control<br>(n=20) | Intervention<br>(n=20) | *p*-value |
| --- | --- | --- | --- |
| **Demographic characteristics** |  |  |  |
| Age (yr) - median (IQR) | 63 (56-67) | 64 (50-73) | 0.93 |
| Male sex - n (%) | 17 (85%) | 15 (75%) | 0.70 |
| **Characteristics of cardiac arrest** |  |  |  |
| Location of cardiac arrest – n (%) |  |  | 1.00 |
| OHCA | 20 (100%) | 19 (95%) |  |
| IHCA | 0 (0%) | 1 (5%) |  |
| Presumed cause of cardiac arrest – n (%) |  |  | 1.00 |
| Cardiac | 19 (95%) | 19 (95%) |  |
| Other | 0 (0%) | 0 (0%) |  |
| Unknown | 1 (5%) | 1 (5%) |  |
| Bystander-witnessed cardiac arrest – n (%) | 18 (90%) | 18 (90%) | 1.00 |
| First monitored rhythm – n (%) |  |  | 0.39 |
| VF | 17 (85%) | 18 (90%) |  |
| VT | 0 (0%) | 1 (5%) |  |
| PAE | 1 (5%) | 1 (5%) |  |
| Unknown | 2 (10%) | 0 (0%) |  |
| Time (min) from cardiac arrest to – median (IQR) |  |  |  |
| start basic life support | 0.5 (0.0-2.8) | 1.0 (0.0-3.3) | 0.80 |
| return of spontaneous circulation | 10 (6-14) | 11 (8-13) | 0.63 |
| **Clinical characteristics at hospital admission** |  |  |  |
| APACHE IV score – median (IQR) | 77 (64-104) | 90 (67-99) | 0.72 |
| Total Glasgow coma scale – median (IQR) | 3 (3-6) | 3 (3-5) | 0.51 |
| Pupil Reflex – n (%) |  |  | 0.18 |
| Bilateral present | 12 (60%) | 17 (85%) |  |
| Bilateral absent | 1 (5%) | 0 (0%) |  |
| Unknown | 7 (35%) | 3 (15%) |  |
| **Sedation** |  |  |  |
| Use of sedation and analgesia – n (%) |  |  |  |
| Propofol | 20 (100%) | 20 (100 %) | - |
| Midazolam | 1 (5%) | 0 (0%) | 1.00 |
| Dexmedetomidine | 1 (5%) | 0 (0%) | 1.00 |
| Morphine | 4 (20%) | 9 (45%) | 0.18 |
| Fentanyl | 13 (65%) | 9 (45%) | 0.34 |
| Remifentanyl | 1 (5 %) | 0 (0%) | 1.00 |

**Figure 1:**
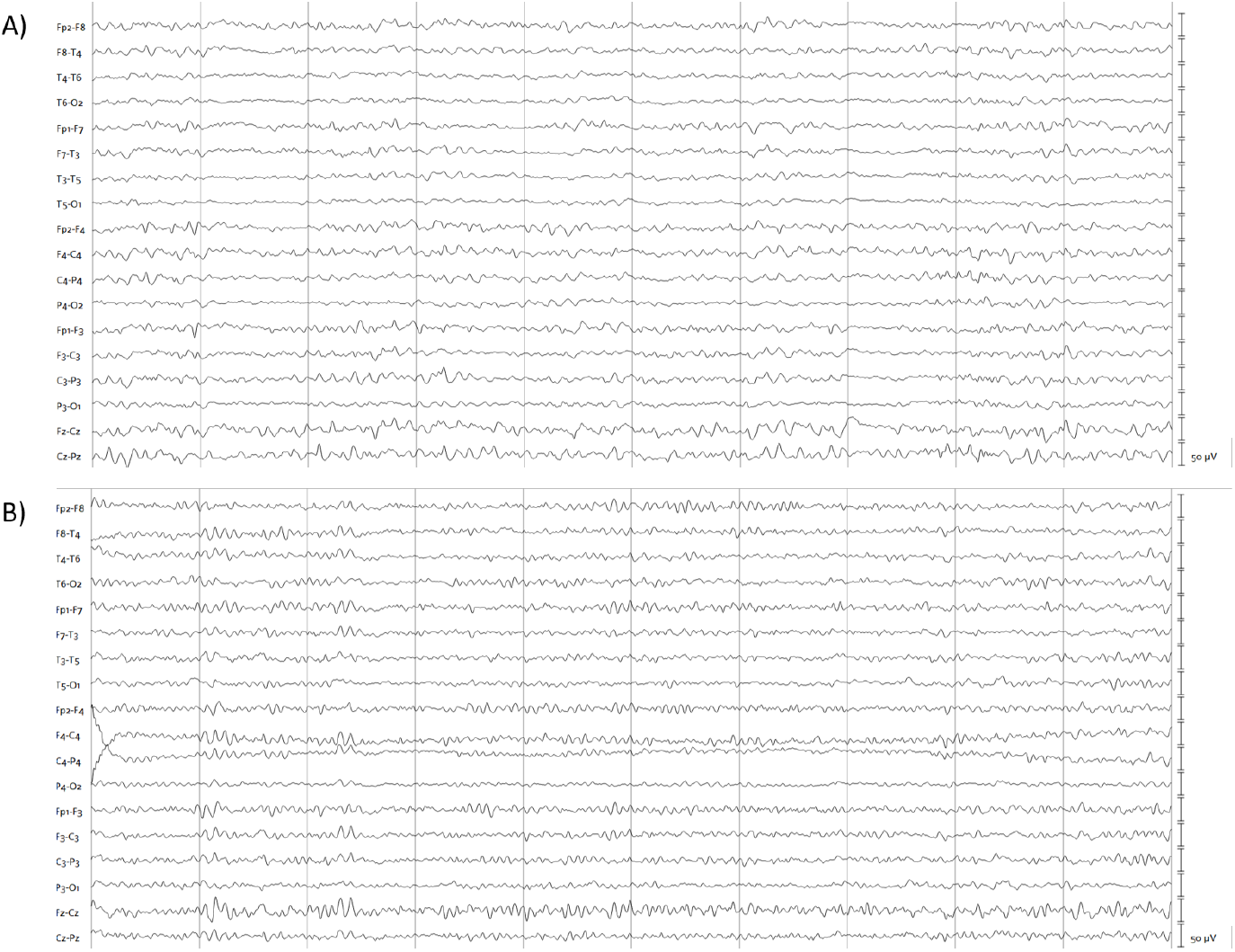
Fragments of EEG recordings from two study patients obtained 3.5 h and 5 h after cardiac arrest, respectively. The EEG was band-pass filtered (0.5–35 Hz). The interval between the two grey vertical lines is 1 s.

### 3.2. Feasibility

The duration of mechanical ventilation was significantly shorter in the intervention group than in the control group (median 12 vs 28 h, *p <* 0.001). ICU length of stay and total sedation time were also significantly shorter (≥ 50%) in the intervention group (see Table 2). These findings indicate that early withdrawal of sedation was feasible.

**Table 2:** Primary and secondary feasibility outcome measures.

|  | **Control**<br>**(n=20)** | **Intervention**<br>**(n=20)** | ***p*-value** |
| --- | --- | --- | --- |
|  | **Median (IQR)** | **Median (IQR)** |  |
| Duration of mechanical ventilation (h) | 28 (22-48) | 12 (7-20) | <0.001 |
| Length of ICU stay (days) | 2.5 (1.9-7.3) | 1.2 (0.9-2.1) | 0.001 |
| Total sedation time (h) | 27 (22-39) | 12 (7-20) | <0.001 |

### 3.3. Safety

There was no increase in the number of complications during ICU stay in the intervention group compared with the control group (Table 3), indicating that early withdrawal of sedation was safe.

**Table 3:** Complications during ICU admission.

|  | **Control**<br>**(n=20)** | **Intervention**<br>**(n=20)** | **p-value** |
| --- | --- | --- | --- |
|  | **Number of**<br>**patients with** | **Number of**<br>**patients with** |  |
| Need for restarting sedation | 7 | 7 | 1.00 |
| Need for re-intubation | 3 | 1 | 0.61 |
| Autodetubation | 0 | 1<sup>a</sup> | 1.00 |
| Death | 0 | 0 | - |
| Cardiac arrhythmia | 4<sup>b</sup> | 1<sup>c</sup> | 0.34 |
| New cardiac arrest | 2 | 0 | 0.49 |
| Pneumonia | 2 | 1 | 1.00 |
| Sepsis | 2 | 0 | 0.49 |
| Bleeding | 0 | 0 | - |
| Delirium | 3 | 1 | 0.60 |
| Haloperidol administered | 2 | 1 | 1.00 |
| Thrombopenia | 1 | 1 | 1.00 |
| Other SAEs during ICU stay | 0 | 1<sup>d</sup> | 1.00 |
| SAEs during rest of hospital stay | 0 | 1<sup>c</sup> | 1.00 |

<sup>a</sup> During preparation for extubation, one patient in the intervention group self-extubated.
<sup>b</sup> Four patients in the control group experienced cardiac arrhythmias during ICU stay: one with 37 episodes of ventricular fibrillation (VF) or ventricular tachycardia (VT) and multiple cardiac arrests over several days; one with slow VT; one with non-sustained VT; and one with VF resulting in cardiac arrest.
<sup>c</sup> One patient in the intervention group experienced one episode of bradycardia (40 bpm) and one episode of non-sustained VT during ICU stay. After transfer to the cardiology department, the patient had another episode of VT resulting in an in-hospital cardiac arrest. The event was witnessed, and basic life support was immediately initiated, with return of spontaneous circulation after a single defibrillation. The patient recovered well from this event.
<sup>d</sup> One patient in the intervention group experienced endotracheal tube dislocation and cuff leakage due to coughing and restlessness after cessation of sedation. Sedation was restarted, and the tube was exchanged without complications.

### 3.4. Neurological outcome

All patients survived the 3- and 6-month follow-up periods. Follow-up was successful in 37 patients (95%) at both time points. All patients had a CPC of 1 or 2, while GOSE scores ranged from 4 to 8 (Figure 2). No statistically significant differences were observed between the intervention and control groups in CPC, GOSE, or Telephone Montreal Cognitive Assessment (T-MoCA) scores at either 3 or 6 months.

**Figure 2:**
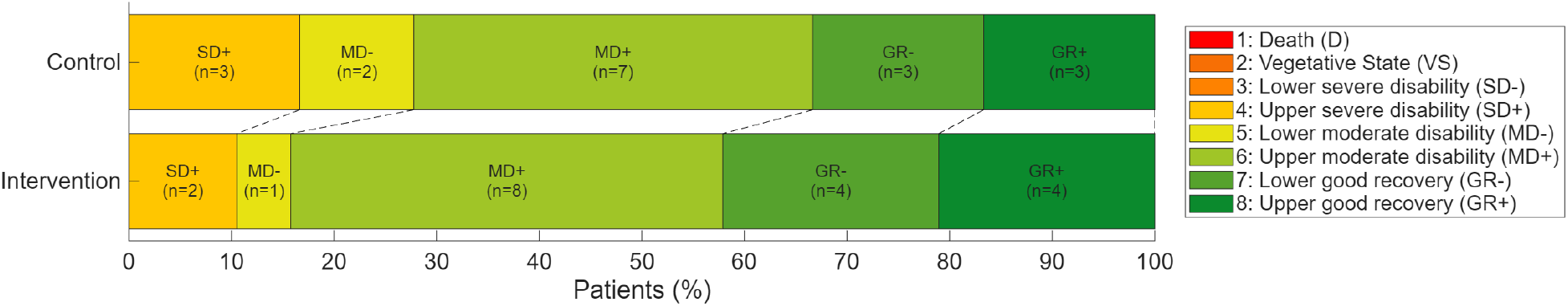
Glasgow Outcome Scale–Extended (GOSE) at 6 months. Patients in the control group received standard care, including deep sedation, targeted temperature management (TTM), and mechanical ventilation for at least 24–48 hours. Patients in the intervention group underwent early cessation of sedation and TTM as soon as clinically feasible after detection of a favourable EEG pattern, followed by weaning from mechanical ventilation. GOSE scores range from 1 to 8, with 1 indicating death and 8 indicating no neurological disability. All patients in both groups had a GOSE score of 4 or higher. No statistically significant differences were observed between the groups; however, where differences were present, outcomes tended to be slightly better in the intervention group.

Detailed results are provided in Table A.4 in the Supplementary Material.

Two patients in the control group experienced a SAE after hospital discharge. One patient was readmitted to another hospital for angina pectoris and was discharged without intervention following evaluation. The second patient was hospitalised after a first seizure that was complicated by a thoracic vertebral fracture. No patients in the intervention group experienced a SAE after hospital discharge.

### 3.5. Patients’ experience with deferred informed consent

All patients rated their experience with deferred consent as neutral or pleasant. On a five-point scale, 3% of the patients rated the procedure as “very pleasant”, 61% as “pleasant,” and 36% as “neither pleasant nor unpleasant”. The majority of patients (97%) indicated that they would not have preferred to be informed about the study at an earlier stage; only one patient (3%) expressed a preference for earlier information.

## 4. Discussion

In this pilot study, early withdrawal of temperature management, sedation, and mechanical ventilation was feasible and safe in patients with a favourable EEG pattern within 12 hours after cardiac arrest. We observed reductions of more than 50% in the duration of mechanical ventilation, sedation, and ICU stay, without an increase in ICU complications or other safety issues. No statistically significant differences in neurological outcomes were observed at 3 or 6 months. However, this safety and feasibility study was not powered to detect differences in neurological outcomes. Even in the absence of improved neurological outcomes, EEG-based weaning strategies may still be clinically valuable. By substantially reducing ICU resource utilisation, such an approach has the potential to lower healthcare costs and promote more appropriate, value-based care.

Previous retrospective studies have suggested that the effect of TTM may depend on the severity of encephalopathy, with patients exhibiting no or only mild encephalopathy having less benefit. A large retrospective study of 1319 comatose patients after cardiac arrest from the University of Pittsburgh demonstrated a potential benefit of TTM at 33 °C compared with 36 °C in patients with the most severe post–cardiac arrest illness, provided that severe cerebral edema or malignant EEG patterns were absent [7]. In contrast, among patients with lower illness severity, treatment at 36 °C was associated with more favourable outcomes. Similarly, a recent post hoc analysis of a prospective cohort study including 479 patients showed that the effect of TTM was associated with encephalopathy severity [8]. Specifically, no significant differences in outcomes between TTM at 33 °C and 36 °C were observed in patients with mild encephalopathy, as indicated by an early favourable EEG. Together, these findings support the concept of EEG-guided, severity-adapted post–cardiac arrest care.

Notably, most recent large TTM trials and ongoing trials evaluating less intensive post–cardiac arrest care (e.g., reduced sedation and device-based fever management, such as STEPCARE) have not routinely incorporated encephalopathy severity (e.g., early EEG) as a stratification variable at randomisation. This may have diluted potential treatment effects in subgroups [4, 1, 12, 13].

Over the past decades, there has been a clear shift from deep sedation and TTM at 33 °C, as applied in the early TTM trials [14, 15], toward less intensive ICU management, including higher temperature targets (36 °C) or even normothermia [4, 1]. The ongoing STEPCARE trial will further study this paradigm by evaluating the effects of no TTM (no early fever management with temperature management devices) and reduced sedation. Our pilot study fits within this overall trend towards reducing ICU treatment intensity, but differs by applying a selective approach focused on patients with mild encephalopathy, as identified by an early favourable EEG.

EEG largely reflects synchronised postsynaptic potentials in cortical pyramidal neurons and, therefore, provides a direct measure of functional synaptic integrity. Synaptic transmission is among the earliest processes affected by cerebral ischemia, rendering EEG highly sensitive to ischemic brain injury [16, 17]. Early background EEG patterns are robust predictors of long-term neurological outcome after cardiac arrest [3, 2, 10, 11]. Moreover, EEG monitoring is bedside-available and relatively low-cost, supporting its use as a practical biomarker to guide individualised post–cardiac arrest care.

If early withdrawal of sedation does influence neurological outcomes, our findings suggest that any effect is more likely to be beneficial rather than harmful. However, the study was underpowered to find any potential differences in outcome. In addition, patients were not randomised between treatment groups, and follow-up assessments were not blinded in this pilot study. Consequently, no definitive conclusions regarding neurological outcomes can be drawn.

In this study, neurological and cognitive outcomes were assessed using the GOSE, CPC, and T-MoCA scales. Overall, patients demonstrated relatively good neurological outcomes, as expected given their favourable EEG patterns [2]. Nevertheless, many patients reported more subtle neurological complaints, such as reduced concentration and increased fatigue. Cognitive impairments after out-of-hospital cardiac arrest are common, affecting up to 50% of patients[18]. These issues are not always captured by the ordinal GOSE and CPC scales.

In conclusion, our study demonstrates that early withdrawal of sedation, TTM, and mechanical ventilation is both feasible and safe in patients with favourable EEG patterns within 12 hours after cardiac arrest. This approach resulted in more than 50% reduction in the duration of mechanical ventilation, sedation, and ICU stay, without an associated increase in complications. Larger studies are needed to evaluate the impact on long-term neurological recovery.

## Data Availability

All data produced in the present study are available upon reasonable request to the authors

## Acknowledgements

We thank all neurologists, lab technicians, intensive care physicians and research nurses from MST and Rijnstate for their participation in patient recruitment and data collection. We also thank the members of the DSMB for their valuable support and their critical review of the study.

JH is supported by a clinical established investigator grant of the Dutch Heart Foundation (Grant Number, 2018T070).

## 5. Ethical Publication Statement

We confirm that we have read the Journal’s position on issues involved in ethical publication and affirm that this report is consistent with those guidelines.

## Appendix A

Detailed information on neurological outcome at 3 and 6 months is given in Table A.4.

**Table A.4:** Neurological outcome at 3 and 6 months.

|  | Follow-up at 3 months |  | Follow-up at 6 months |  |
| --- | --- | --- | --- | --- |
|  | Control<br>(n=20) | Intervention<br>(n=20) | Control<br>(n=20) | Intervention<br>(n=20) |
| Mortality - n | 0 | 0 | 0 | 0 |
| GOSE - n |  |  |  |  |
| 1: Dead | 0 | 0 | 0 | 0 |
| 2: Vegetative state | 0 | 0 | 0 | 0 |
| 3: Lower severe disability | 0 | 0 | 0 | 0 |
| 4: Upper severe disability | 5 | 5 | 3 | 2 |
| 5: Lower moderate disability | 1 | 0 | 2 | 1 |
| 6: Upper moderate disability | 5 | 5 | 7 | 8 |
| 7: Lower good recovery | 5 | 4 | 3 | 4 |
| 8: Upper good recovery | 3 | 4 | 3 | 4 |
| Missing | 1 | 2 | 2 | 1 |
| CPC Score - n |  |  |  |  |
| 1: Good cerebral performance | 5 | 11 | 6 | 12 |
| 2: Moderate cerebral disability | 14 | 8 | 13 | 7 |
| 3: Severe cerebral disability | 0 | 0 | 0 | 0 |
| 4: Coma or vegetative state | 0 | 0 | 0 | 0 |
| 5: Brain death | 0 | 0 | 0 | 0 |
| Missing | 1 | 1 | 1 | 1 |
| T-MOCA - Median (IQR) | 20 (19-21) | 19 (18-20) | 21 (20-22) | 21 (20-21) |
| SAE after hospital discharge - n | 1 | 2 | 0 | 0 |

GOSE = Glasgow Outcome Scale Extended, CPC = Cerebral Performance Category, T-MoCA = Montreal Cognitive Assessment by telephone. For this telephone-based assessment, the first two items of the original MoCA cannot be administered, resulting in a maximum score of 22 points.

